# COVID-19 Test Strategy to Guide Quarantine Interval in University Student

**DOI:** 10.1101/2020.12.09.20246785

**Authors:** Jill M. Kolesar, Tyler Gayheart, Lance Poston, Eric Monday, Derek Forster, Elizabeth Belcher, Rani Jaiswal, J. Kirsten Turner, Donna K. Arnett, Eric B. Durbin, Joseph Monroe, Frank Romanelli, Susanne M. Arnold, C. Darrell Jennings, Heidi Weiss, Robert DiPaola

**Affiliations:** College of Pharmacy, University of Kentucky; College of Medicine, University of Kentucky; Markey Cancer Center, University of Kentucky; Information Technology Services, University of Kentucky; Student Success, University of Kentucky; Executive Vice President for Finance and Administration, University of Kentucky; College of Public Health, University of Kentucky; Police Department, University of Kentucky

## Abstract

**BACKGROUND:** Following COVID-19 exposure, the CDC recommends a 10-14 day quarantine for asymptomatic individuals and more recently a 7 day quarantine with a negative PCR test. We performed a university-based prospective student cohort study to determine if early PCR negativity predicts day 14 negativity.

**METHODS:** We enrolled 101 asymptomatic, quarantining, students, performed nasopharyngeal swabs for viral testing on days 3 or 4, 5, 7, 10 and 14 and determined the proportion of concordant negative results for each day versus day 14 with a two-sided 95% exact binomial confidence interval.

**RESULTS:** Overall, 14 of 90 (16%, 95% CI: 9% - 25%) tested positive while in quarantine, with 7 initial positive tests on day 3 or 4, 5 on day 5, 2 on day 7, and none on day 10 or 14. Rates of concordant negative test results are: day 5 vs. day 14 = 45/50 (90%, 95% CI: 78% - 97%); day 7 vs. day 14 = 47/52 (90%, 95% CI: 79% - 97%); day 10 vs. day 14 = 48/53 (91%, 95% CI:79% - 97%), with no evidence of different negative rates between earlier days and day 14 by McNemar’s test, p > 0.05.

**CONCLUSIONS:** The 16% positive rate supports the ongoing need to quarantine close contacts of COVID-19 cases, but this prospective study provides the first direct evidence that exposed asymptomatic students ages 18-44 years in a university setting are at low risk if released from quarantine at 7 days if they test negative PCR test prior to release.

---

Coronavirus disease 2019 (COVID-19) is an infectious disease caused by the novel coronavirus SARS-CoV-2, first reported in Wuhan, China on December 31, 2019.^1^ The disease was declared a global emergency by the World Health Organization on January 30, 2020 and a global pandemic on March 11, 2020.^2^ COVID-19 is primarily spread by respiratory droplets, with an estimated reproduction number (R_0_) of 2.8-5.5,^3^ which suggests COVID-19 is more transmissible than seasonal influenza, with an R_0_ of 1.8.^4^ While the majority of individuals with a symptomatic infection have mild symptoms, the case fatality rate is approximately 1%,^1,2^ making COVID-19 more pathogenic than seasonal influenza pandemics with a case fatality rate of approximately 0.01%.^5^

Since effective pharmacological interventions are limited, public health measures including social distancing, isolation and quarantine are employed to limit spread of the virus, reduce infections and prevent deaths. Isolation separates infected individuals and quarantine separates asymptomatic individuals who have had contact with a COVID-19 case..^6^ Currently, there are no prospective studies demonstrating the benefit of quarantine to prevent COVID-19 infections. A recent Cochrane review includes modeling studies that consistently suggest isolation of infected individuals and quarantine of contacts is beneficial, with 44-88% of cases averted and 31-63% of deaths, although the authors rated the evidence of certainty as low.^7^ Additional prospective studies, especially in University settings, are warranted to so that informed decisions can be with regard to quarantine status of students.

The CDC currently recommends a 14 day quarantine^6^ based on an early report from WHO-China, suggesting a mean viral incubation period of 5-6 days with a range of 14 days.^1^ Recently, among asymptomatic individuals a 10 day quarantine, or a 7 day quarantine with a negative PCR test was determined to be acceptable by the CDC.^6^ Rather than relying on prospective studies, this recommendation is based on modeling studies (pending peer review),^8,9^ showing the post-quarantine transmission risk was approximately 4% at 7 days with a negative test and 1.4% at 10 days without testing. Given the limited data supporting these recommendations, the need to understand the applicability for students in the university setting, and the negative financial, emotional and logistical impact of an extended quarantine period,^10^ the purpose of this study was to test the hypothesis that consistent testing throughout the quarantine period might reliably predict day 14 outcome and therefore could be used to determine if asymptomatic students might qualify for a shortened quarantine duration.

## METHODS

### STUDY DESIGN

START is a prospective cohort study. Quarantined students were enrolled between September 24, 2020 and November 24, 2020, with the last student completing their 14-day quarantine on December 6, 2020. Per the IRB-approved protocol (NCT04573634), each student who entered quarantine was invited to participate in the study. Individuals who elected to participate were called by study personnel and the consent process was performed over the telephone with an emailed consent document. At study entry, concierge testing was scheduled by UK HealthCorps and conducted by an outside vendor who went to the student’s location. Test results were transmitted to UK HealthCorps. Students received a $10 gift card for each PCR test completed which was emailed to them after they completed the study.

Age, gender, symptom status and the outcome of the viral PCR test were provided to the study team by UK HealthCorps. Participants were emailed a survey to characterize COVID-19 exposures, including length of exposure, whether the exposure was a household contact, and whether there was direct physical contact or sharing of secretions. Surveys were initially administered at enrollment and non-responsive individuals received up to two additional attempts spaced 2 weeks apart.

### STUDY POPULATION

Participants were students at the University of Kentucky who were >18 years of age, asymptomatic and quarantined for 14 days due to a close contact with a known COVID-19 case. Participants were required to enroll within 3 days of exposure but were not required to have a negative test at study entry. Participants were excluded from the study population if they had a psychiatric illness or social situation that would limit compliance with study requirements.

### TESTING

SARS-CoV2 NAAT testing was performed by a commercial vendor (WildHealth) utilizing a laboratory developed qPCR test on nasopharyngeal swab on days 3 or 4, 5, 7, 10 and 14 of the quarantine period. Nucleic acid was extracted and purified from the sample using an automated system (Kingfisher, ThermoFisher Scientific). Two regions of the SARS-CoV2 N-gene specific RNA was identified by LDT specific primers and amplified (TaqPath™ 1-Step RT-qPCR Master Mix) and detected using a quantitative RT-PCR reaction (QuantStudio 5, ThermoFisher Scientific).

### OVERSIGHT AND FUNDING

The study was registered at ClinicalTrials.gov (NCT04573634) and approved by the University of Kentucky IRB (UK #59793). Study conduct was in accordance with the principles of the Declaration of Helsinki and Good Clinical Practice guidelines. The only exclusions from the analyses were three individuals who were ineligible because they refused to quarantine. Gift cards were provided by the University of Kentucky COVID-19 response team and the Markey Cancer Center supported the statistical analysis.

### OUTCOMES

The START study opened on June 17, 2020 with the primary endpoint to determine the penetrance of COVID-19 in Kentucky by assessing the prevalence of IgG antibodies to SARS CoV-2. It was amended on September 18, 2020 to add a student quarantine cohort and two secondary endpoints were added. 1) determine PCR positivity over a 14-day quarantine period. 2) determine if PCR negativity on days 3, 5, 7 or 10 by standard of care testing predicts PCR negativity on day 14.

### SAMPLE SIZE

The main goal of the study was to estimate the proportion of concordant negative test results from earlier days vs. day 14. The planned total sample size of 100 students completing a 14-day quarantine provides an estimate of overall positivity rate, rates on specific days of quarantine and the proportion of concordant negative results for each day versus day 14 with a two-sided 95% exact binomial confidence interval with a width that is no larger than 30%.

### STATISTICAL ANALYSIS

Descriptive statistics were calculated to summarize demographic and student exposure characteristics. The proportions of concordant negative test results between days 5, 7 and 10 versus day 14 were estimated along with two-sided 95% exact binomial confidence intervals. A McNemar’s test for these paired time point results was utilized to compare the proportion of negative results on earlier days versus day 14.

## RESULTS

Between 9.24.2020 and 11.24.2020, a total of 741 students were invited to participate; 331 declined participation, 324 were unreachable, 3 refused to quarantine and 101 students enrolled. **See Figure 1**. After enrollment, two students refused additional testing and withdrew. Age, gender and symptoms status are available for 99 students. **See Table 1**. Nine of these students did not formally withdraw but did not complete the survey or testing. The median age of study participants was 21 (range 18-44) years. There were 68 female (69%) and 31 male (31%) participants, with 33/99 (33%) being exposed to an asymptomatic individual, 43/99 (43%) being exposed to a symptomatic individual and 23/99 (23%) not knowing the status of their contact.

**Figure 1:**
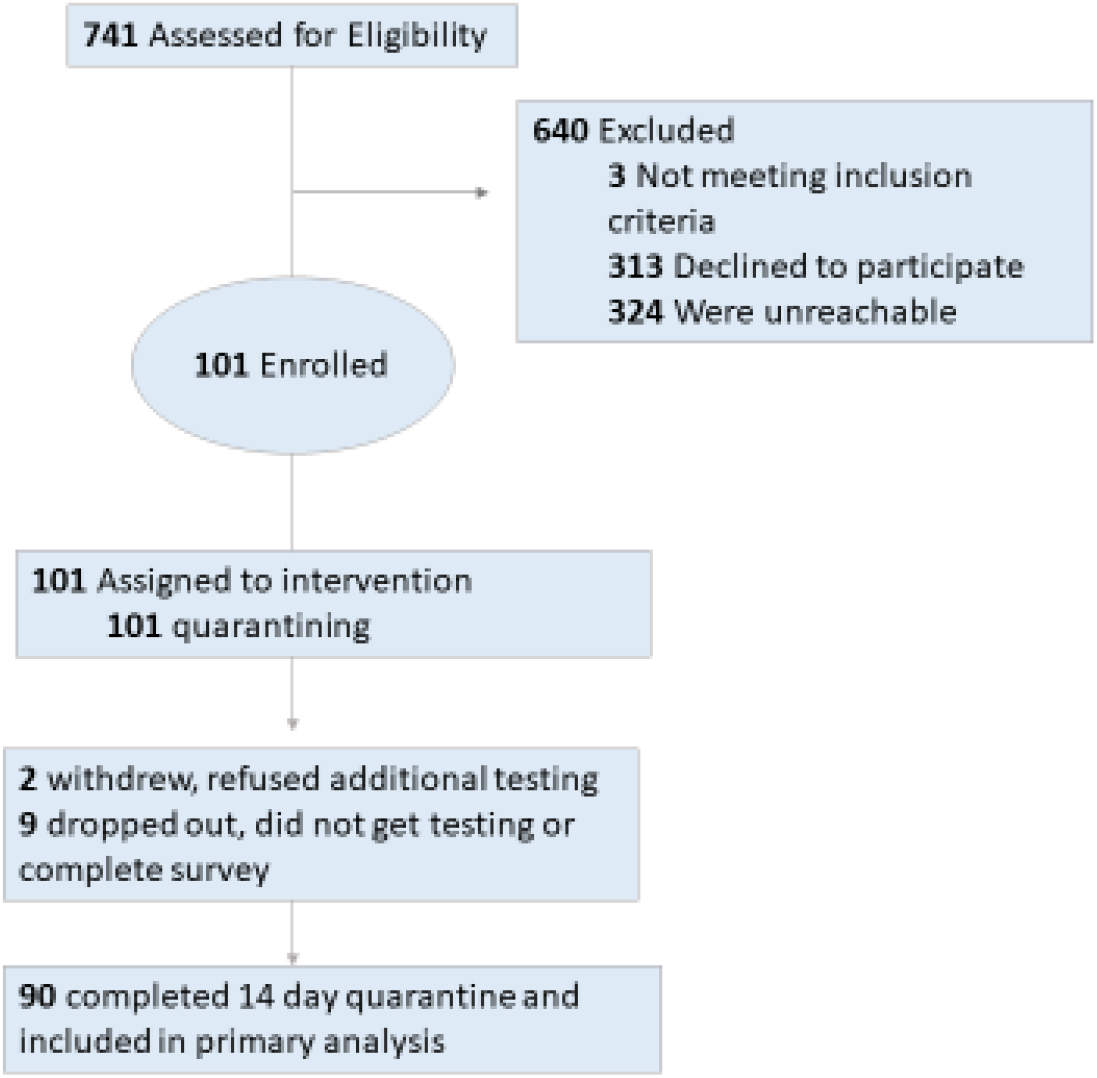
Subject Enrollment.

**Table 1:** Characteristics of Study Participants.

|  |  |  | Ever<br>COVID-19<br>Positive | Remained<br>COVID-19<br>Negative | No COVID-19<sup>&</sup><br>Result |
| --- | --- | --- | --- | --- | --- |
| **Gender** |  |  |  |  |  |
|  | Female | 68 | 10 | 52 | 6 |
|  | Male | 31 | 4 | 23 | 4 |
| **Age** |  |  |  |  |  |
|  | Median | 21 |  |  |  |
|  | Range | 18-44 |  |  |  |
| **Exposure to Symptomatic Individual** |  |  |  |  |  |
|  | No | 33 | 1 (3%) | 27 (82%) | 5 (15%) |
|  | Yes | 43 | 7 (16%) | 32 (74%) | 4 (9%) |
|  | Status Not Known | 23 | 6 (26%) | 16 (70%) | 1 (4%) |
| **Household Contact** |  |  |  |  |  |
|  | Yes | 39 | 4 (10%) | 31 (80%) | 4 (10%) |
|  | No | 41 | 7 (17%) | 31 (76%) | 3 (7%) |
|  | Missing data* | 19 | 3 (16%) | 13 (68%) | 3 (16%) |
| **Unmasked Exposure Time** |  |  |  |  |  |
|  | < 1 hr | 18 | 1 (6%) | 17 (94%) | 0 |
|  | 1-3 hr | 30 | 5 (17%) | 20 (67%) | 5 (17%) |
|  | >3 hr | 32 | 5 (16%) | 24 (75%) | 2 (6%) |
|  | Missing data* | 19 | 3 (16%) | 13 (68%) | 3 (16%) |
| **Direct Physical Contact** |  |  |  |  |  |
|  | Yes | 24 | 3 (13%) | 19 (79%) | 2 (8%) |
|  | No | 56 | 8 (14%) | 43 (77%) | 5 (9%) |
|  | Missing data* | 19 | 3 (16%) | 13 (68%) | 3 (16%) |
| **Exchange of Secretions** |  |  |  |  |  |
|  | Yes | 18 | 3 (17%) | 14 (77%) | 1 (6%) |
|  | No | 62 | 8 (13%) | 48 (77%) | 6 (10%) |
|  | Missing data* | 19 | 3 (16%) | 13 (68%) | 3 (16%) |

\*Did not complete survey; <sup>&</sup>Did not have testing

The study population completed a median of 4 PCR tests. Overall, 14 of 90 (16%) had at least one positive test while in quarantine. Of the 14 students testing positive, 7 individuals tested positive initially on day 3 or 4, 5 individuals on day 5, and 2 individuals on day 7. No students tested positive initially on day 10 or 14. The rates of concordant negative test results among those who completed tests from earlier days and day 14 are as follows: day 5 vs. day 14 = 45/50 (90% concordant negative, 95% CI: 78% - 97%); day 7 vs. day 14 = 47/52 (90% concordant negative, 95% CI: 79% - 97%); day 10 vs. day 14 = 48/53 (91% concordant negative, 95% CI:79% - 97%). A McNemar’s test for these paired time points did not reach statistical significance (p > 0.05) indicating no evidence of differences in negative rates between earlier days and day 14. There were no instances where a negative test in earlier days was discordant with a positive result on day 14. Being exposed to someone with an asymptomatic infection may be associated with lower risk of transmission, with only 3% (1/33) of these individuals developing an infection, while not knowing the symptom status of the case to which they were exposed appeared to increase the risk with 26% (6/23) developing a COVID infection. Of the other contact measures, only unmasked exposure time appeared to influence the risk of transmission, with only 6% (1/18) of individuals exposed for less than an hour developing an infection.

## DISCUSSION

This is the first prospective study of students in a university setting assessing a testing strategy to predict day 14 positivity to evaluate the safety of shortened quarantine recommendations for young adults. Our study demonstrated that the overall COVID-19 positivity rate in quarantining individuals was 16%. These results are consistent with a recent meta-analysis that evaluated the transmission rate of COVID-19 among household contacts.^11^ Including 40 publications and more than 30,000 individuals, the authors estimated the overall household secondary attack rate (defined as the number of new cases divided by the total number of individuals exposed in the household) to be 18.8% (95% confidence interval [CI]: 15.4%–22.2%).^11^ We observed similar transmission rates in our study for both household and non-household transmission in this study, supporting the need for quarantine after exposure.

Since quarantine times are poorly adhered to, with only 10.9% of quarantining individuals remaining at home in one report,^14^ a testing strategy that allows for a reduction in quarantine time could lead to improved compliance. The 14-day quarantine in asymptomatic individuals is based on the probability of exposed individuals developing symptoms, which is 50% at 5 days^12^ and > 95% at 14.^13^ In this study, all quarantining students who developed a COVID-19 infection were positive by day 7 post exposure. Concordance of negativity rates between day 5 and later dates with day 14 were consistently high, ranging from 90% to 91%. Consistent with CDC recommendations,^6^ this study provides prospective evidence that a negative COVID-19 test in young adults on day 7 predicts those unlikely to develop a COVID-19 infection and supports the CDC recommendation that 7 days with a negative test may be an adequate quarantine period in the university setting.

There are approximately 20 million students enrolled in institutions of higher education in the United States. Mental health issues are common among college students, with 25% being treated for anxiety or depression and suicide the second leading cause of death in this population.^15^ When institutions of higher education converted to almost complete remote, on-line instruction in response to the COVID-19 pandemic, usual college activities that foster engagement and belonging were also suspended, resulting in increased stress and anxiety in most individuals and many concerns related to decreased social interactions due to physical distancing. Isolation, including during a prolonged quarantine, may have significant negative impacts on college students’ emotional health as well as the overall vitality of colleges and universities.^16,17^ Our results suggest that the quarantine period could safely be reduced to 7 days for college-age students.

Strengths of this study include its prospective design with a priori sampling size requirements designed to estimate rates with a specified level of precision. The resulting concordant negative rate estimates and level of precision were consistent and are similar across all timepoints. The primary limitation is a potential lack of generalizability as it was conducted in a relatively young and asymptomatic population in a single geographic university setting and testing was performed by nasopharangeal swab. Whether these findings apply to a wider population or other testing methods are not known. In addition, we had a relatively large drop-out rate (11/101, 11%). Despite this, our estimates are precise and reliable and provide prospective confirmation of the current modeling estimates that suggest a 7day quarantine time is adequate in asymptomatic low-risk individuals. This study contributes important data to support a critically needed data-driven approach to COVID-19 related decision making.^15^ Finally, as noted in CDC guidelines, local public health departments guide local quarantine recommendations and decisions should be based on understanding and balancing the potential risk of shorter quarantine periods and consideration of local circumstances in this evolving pandemic.

## Data Availability

The data will be available by emailing

## Acknowledgements

The study investigators gratefully acknowledge the quarantined students who made this study possible, study staff, Justin Levens who managed the data and Eric Orr, who coordinated between the study team and the University of Kentucky (UK) HealthCorps. The authors also acknowledge the UK Screening Testing and Tracing to Accelerate Restart and Transition (START) team for helpful discussion and critical review of the manuscript, the START team includes; Robert DiPaola, MD, Donna K. Arnett, PhD, Susanne Arnold, MD, Jay Blanton, Richard Chapman, Becky Dutch, PhD, Derek Forster, MD, Tyler Gayheart, PhD, Lauren Greathouse, Hanine El Haddad, MD, Cliff Iler, JD, C. Darrel Jennings, MD, Jill Kolesar, PharmD, Ian McClure, JD, Erin McMahon, JD, Brian Nichols, Lance Poston, PhD, Evan Ramsay, Frank Romanelli, PharmD, Jennifer Rose, Mathew Sanger, Colleen Swartz, DNP, Heidi Weiss, PhD, Pamela Woods.

## Notes

### Competing Interest Statement

The authors have declared no competing interest.

### Clinical Trial

NCT04573634

### Funding Statement

NCI Cancer Center Support Grant (P30 CA177558)
Gift cards were provided by the University of Kentucky COVID-19 response team and the Markey Cancer Center supported the statistical analysis.

### Author Declarations

The study was registered at ClinicalTrials.gov (NCT04573634) and approved by the University of Kentucky IRB (UK #59793). Study conduct was in accordance with the principles of the Declaration of Helsinki and Good Clinical Practice guidelines.

## References

1. World Health Organization. Report of the WHO-China Joint Mission on Coronavirus Disease 2019 (COVID-19). 24 Feb 2020. https://www.who.int/docs/default-source/coronaviruse/who-china-joint-mission-oncovid-19-final-report.pdf. Accessed November 24, 2020

2. World Health Organization (WHO). Rolling updates on coronavirus diseases (COVID-19). www.who.int/emergencies/diseases/novel-coronavirus-2019/events-as-they-happen Accessed 8 December 2020).

3. Liu Y, Gayle AA, Wilder-Smith A, Rocklov J. The reproductive number of COVID-19 is higher compared to SARS coronavirus. J Travel Med; 2020 Mar 13;27(2):taaa021.

4. Biggerstaff M, Cauchemez S, Reed C, Gambhir M, Finelli L. Estimates of the reproduction number for seasonal, pandemic, and zoonotic influenza: a systematic review of the literature. BMC Infect Dis 2014 Sep 4;14:480. doi: 10.1186/1471-2334-14-480

5. Taubenberger JK, Morens DM. 1918 Influenza: the mother of all pandemics. Emerg Infect Dis 2006 Jan;12(1):15-22.

6. Centers for Disease Control and Prevention. Coronavirus Disease 2019. https://www.cdc.gov/coronavirus/2019-ncov/more/scientific-brief-options-to-reduce-quarantine.html Accessed December 8, 2020

7. Nussbaumer-Streit B, Mayr V, Dobrescu AI, Chapman A, Persad E, Klerings I, Wagner G, Siebert U, Christof C, Zachariah C, Gartlehner G. Quarantine alone or in combination with other public health measures to control COVID-19: a rapid review. Cochrane Database Syst Rev. 2020 Apr 8;4(4):CD013574.

8. Quilty BJ, Clifford S, Flasche S, Kucharski AJ, CMMID COVID-19 Working Group, Edmunds WJ. Quarantine and testing strategies in contact tracing for SARS-CoV-2: a modelling study. medRxiv. 2020. https://doi.org/10.1101/2020.08.21.20177808

9. Wells CR, Townsend JP, Pandey A, et al. Optimal COVID-19 quarantine and testing strategies. medRxiv. 2020.10.1101/2020.10.27.20211631. https://doi.org/10.1101/2020.10.27.20211631e

10. Singh SP, Khokhar A. Prevalence of Posttraumatic Stress Disorder and Depression in General Population in India During COVID-19 Pandemic Home Quarantine. Asia Pac J Public Health. 2020 Nov 16:1010539520968455.

11. Madewell JZ, Yang Y, Longini IM, Halloran EM, Dean NE. Household transmission of SARS-CoV-2: a systematic review and meta-analysis of secondary attack rate medRxiv 2020 Jul 31;2020.07.29.20164590.

12. Kucirka LM, Lauer SA, Laeyendecker O, Boon D, Lessler J. Variation in False-Negative Rate of ReverseTranscriptase Polymerase Chain Reaction–Based SARS-CoV-2 Tests by Time Since Exposure. Ann Intern Med. 2020 May 13;M20-1495.

13. Ashcroft P, Huisman JS, Lehtinen S, Bouman JA, Althaus CL, Regoes RR, et al. COVID-19 infectivity profile correction. Swiss Med Wkly [Internet]. 2020 Aug 5 [cited 2020 Sep 11];150(3132). Available from: https://smw.ch/article/doi/smw.2020.20336

14. Smith LE, Potts HWW, Amlot R, Fear NT, Michie S, Rubin J. Adherence to the test, trace and isolatesystem: results from a time series of 21 nationally representative surveys in the UK (the COVID-19 RapidSurvey of Adherence to Interventions and Responses [CORSAIR] study) [Internet]. Public and GlobalHealth; 2020 Sep [cited 2020 Sep 30]. Available from:http://medrxiv.org/lookup/doi/10.1101/2020.09.15.2019

15. Lederer AM, Hoban MT, Lipson SK, Zhou S, Eisenberg D. More Than Inconvenienced: The Unique Needs of U.S. College Students During the COVID-19 Pandemic. Health Educ Behav. 2020 Oct 31:1090198120969372.

16. Wathelet M, Duhem S, Vaiva G, et al. Factors associated with mental health disorders among university students in France confined during the COVID-19 pandemic. JAMA Netw Open. 2020;3(10):e2025591.

17. Son C, Hegde S, Smith A, Wang X, Sasangohar F. Effects of COVID-19 on College Students’ Mental Health in the United States: Interview Survey Study. J Med Internet Res. 2020 Sep; 22(9): e21279.

